# Functional Movement Disorder Is Associated with Abnormal Interoceptive Brain Activity: A Task-based Functional MRI Study

**DOI:** 10.1101/2024.07.23.24310881

**Authors:** Primavera A Spagnolo, Jacob A Parker, Mark Hallett, Silvina Horovitz

**Affiliations:** Mary Horrigan Connors Center for Women’s Health and Gender Biology, Brigham and Women’s Hospital, Boston, MA, USA; Department of Psychiatry, Brigham and Women’s Hospital, Boston, MA, USA; Harvard Medical School, Boston, MA, USA; Neuroscience Graduate Group; University of Pennsylvania, Philadelphia, PA, USA; Human Motor Control Section, Medical Neurology Branch, National Institute of Neurological Disorders and Stroke, National Institutes of Health, Building 10, Room 7D37, 10 Center Drive, Bethesda, MD 20892-1428, USA; National Institute of Neurological Disorders and Stroke, National Institutes of Health, Bethesda, Maryland, USA

**Keywords:** Functional Movement Disorder, Interoception, Prediction Error, Insula, Default Mode Network

## Abstract

**Background:** Aberrant interoceptive processing has been hypothesized to contribute to the pathophysiology of functional neurological disorder, although findings have been inconsistent. Here, we utilized functional magnetic resonance imaging (fMRI) to examine neural correlates of interoceptive attention – the conscious focus and awareness of bodily sensations – in functional movement disorder (FMD).

**Methods:** We used voxelwise analyses to compare blood oxygenation level-dependent responses between 13 adults with hyperkinetic FMD and 13 healthy controls (HCs) during a task requiring attention to different bodily sensations and to an exteroceptive stimulus. Additionally, we examined between-group differences in self-reported measures of interoception and evaluated their relationship with neural activity.

**Results:** Interoceptive conditions (heartbeat, stomach and ‘body’, indicating sensations from the body part or limb affected in FMD participants) activated a network involving the precuneus, the posterior cingulate cortex (PCC) and caudate nucleus (CN) bilaterally, and the right anterior insula (aINS) (*p* <0.05 , corrected). Group differences in brain activity were mainly driven by processing of disease-related interoceptive signals, which in the FMD group was associated with a broader neural activation than monitoring gastric interoception, while no group differences were detected during cardiac interoception. Differences based on interoceptive focus (*body vs* heartbeat and stomach) between FMD subjects and HCs were found in PCC, CN, angular gyrus, thalamus, and in the mid-insula (*p* <0.05, corrected).

**Conclusions:** This is, to our knowledge, the first study showing that FMD is associated with abnormal interoceptive processing in regions involved in monitoring body state, attentional focus, and homeostatic inference.

## Introduction

Interoception refers to sensing, interpreting, and integrating a wide range of internal bodily signals^1^. This complex process encompasses different dimensions, including interoceptive accuracy, attention, and sensibility^2,3^, which contribute to the homeostatic regulation of the body as well as to cognition, attention, and emotion processing^4,5^. Furthermore, interoceptive signals are thought to be critical for body awareness and for the generation of subjective motor-related feelings states^6,7^. At the neurocircuitry level, interoceptive processing has been consistently associated with activity in the dorsal mid-insular cortex, as well as in sensorimotor, temporal, and prefrontal cortex regions^8–16^.

Alterations in interoception, and its underlying neurocircuitry, have been increasingly recognized as an important transdiagnostic component of different psychiatric disorders, including anxiety and mood disorders, eating disorders, addictive disorders, and somatic symptom disorders. In recent years, interoceptive deficits have also been proposed to contribute to the generation of functional neurological symptoms^17^. To test this hypothesis, several studies have examined behavioral correlates of interoceptive accuracy (the ability to accurately detect internal bodily sensations) in patients with Functional Neurological Disorders (FND), with a focus on heartbeat perception accuracy^18–25^. Few studies have also examined interoceptive sensibility (subjective perception and beliefs about one’s interoceptive accuracy and attention^23,25,26^ and its correlation with white matter integrity indices in FND patients *vs* healthy controls^27^.

Findings from these studies have been inconsistent, showing either reduced interoceptive accuracy in individuals with functional movement disorder (FMD) and functional seizures (FS)^18,19,22,23,26^ or lack of group-level differences between patients and controls^20,21,25,27^.These discrepancies are likely due to several factors, including potential heterogeneity in interoceptive abilities among patients with different FND subtypes. Furthermore, behavioral measures of cardiac interoception accuracy do not capture impairment in other dimensions of interoception that may be implicated in FND^28^.

In particular, a growing body of evidence suggests that individuals with FND selectively monitor disease-related somatic information and exhibit abnormal body-centered attention. For instance, FND patients have been shown to over-report somatic symptoms, compared to clinical assessment^18^ or objective measures^29^ of symptom frequency. These observations suggest that alterations in interoceptive attention (IA) - the conscious focus and awareness of bodily sensations^28^ – may also contribute to the pathophysiology and symptomatology of FND. However, to date no published study has directly probed IA processing, and its neural correlates, in individuals with FND. Furthermore, research is needed to understand whether IA abilities vary according to the interoceptive signals processed, since it can hypothesized that the increased weight attributed to disease-related interoceptive signals in FND may hijack IA allocated to other bodily signals.

To begin addressing this set of questions, in the current study we investigated neural mechanisms of IA in patients with FMD and healthy controls (HCs) during performance of an interoception attention task comprising different interoceptive conditions and an exteroceptive condition. Furthermore, we examined the relationship between neural activity during IA and individual differences in□self reported interoception, as measured by the Multidimensional Scale of Interoceptive Awareness (MAIA)^30^. We hypothesized that patients with FMD compared to controls would show increased activity in insula during the interoceptive conditions compared to exteroception. We also predicted that the magnitude of the hemodynamic response in this region during IA would correlate with self-reported measures of interoception in patients with FMD.

## Materials and Method

### Subjects

Participants in this study were recruited from the Human Motor Control Clinic at the National Institutes of Health (NIH) between April 2018 and April 2022 and belonged to a larger ongoing study investigating the clinical and neurobiological correlates of FMD. Study subjects partially overlap with those reported in previous articles^31,32^ and included 13 patients with diagnosis of FMD and 13 age- and sex-matched HCs. Exclusion criteria for FMD patients included movement symptoms affecting the head or neck; comorbid neurologic diseases; psychosis, bipolar disorder, or current substance abuse; current suicidality; disease severity requiring hospitalization; use of tricyclic antidepressants or antiepileptic medications; and abnormal clinical MRI brain. HCs were excluded for use of antidepressant medications within the last 6 months.

### Clinical and behavioral assessments

During the study, all participants underwent a physical and neurological exam. Diagnosis of ‘clinically definite’ FMD was made by at least 2 movement disorders specialists utilizing Fahn and Williams criteria. Participants also completed the Simplified-Functional Movement Disorders Rating Scale (S-FMDRS)^33^, which was used to identify the limb or body part most affected, based on symptom severity and frequency. Anxious and depressive symptomatology were evaluated using the Hamilton Anxiety Rating Scale (HAM-A)^34^ and the Hamilton Rating Scale for Depression (HAM-D)^35^. We also administered the Multidimensional Assessment of Interoceptive Awareness (MAIA) to evaluate different self-reported components of interoception^30,36^. This 32-item questionnaire includes eight scales (i.e., Noticing, Not-Distracting, Not-Worrying, Attention Regulation, Emotional Awareness, Self-Regulation, Body Listening, Trusting) and has been extensively employed in both healthy and clinical populations. A major strength of the MAIA is the ability to differentiate between maladaptive and beneficial attention styles towards the body^36^.

As part of the larger study in which they were enrolled, study subjects were also screened for psychiatric diagnoses using the Structured Clinical Interview for DSM-IV-TR, Patient Edition (SCID)^37^.

### Image Acquisition

Imaging was acquired during each visit with a 3-T MR750 GE scanner, using a 32-channel head coil. Each fMRI scan included five consecutive runs in the following order: anatomical scan (∼5 min); resting state (∼6 min), task (3 runs, ∼9 min each), and resting state (∼6 min). A single-shot, multi-echo, echo-planar imaging (EPI) sequence with Sensitivity Encoding (SENSE) was employed for blood oxygenation level-dependent (BOLD) fMRI scans. fMRI acquisition parameters were: repetition time (TR) = 2500 ms, number of echoes = 3, echo times (TEs) = 14.5, 32.3, 50.1 ms, flip angle (FA) = 75°, field of view (FOV) = 216 x 216 mm^2^, matrix size= 72×72, slice thickness: 3.0 mm, slices = 36, nominal voxel size: 3.0 x 3.0 x 3.0 mm^3^, repetitions = 144. For an anatomical reference for the fMRI analyses, a T1-weighted MRI scan with magnetization-prepared rapid gradient echo (MPRAGE) sequence with SENSE was obtained (TR = 7.7ms, TE = 3.436ms, FA = 7°, FOV = 256 x 256 mm^2^, nominal voxel size: 1.0 x 1.0 x 1.0 mm^3^, slices = 176).

### Interoceptive Attention Task

This task was modified from prior fMRI studies of interoception where subjects were asked to attend to their body sensations^8,11,12,14,15,38^. In brief, during each of the 3 runs of the task, participants alternated between two experimental conditions, the interoceptive attention and the exteroceptive attention condition. During the interoception condition, the word “HEART”, “STOMACH”, or “BODY” was presented for 10 seconds in the center of a screen, in black font against a white background. During this time, subjects were instructed to focus their attention on their heartbeat (HB) or stomach distension (S). When patients with FMD saw the word “BODY” on the screen, they were asked to monitor sensations coming from the affected limb/body part (e.g., left leg; right arm), as identified by the S-FMDRS. If patients reported functional motor symptoms in different limbs/body parts, they were instructed to focus on the most severely affected. During these blocks, healthy controls focused on sensations from the limb/body part indicated by the matching FMD patient.

In this version of the task, the interoceptive conditions included monitoring both visceral and somatic signals, in line with the conceptualization of interoception as the sensing of all physiological tissues that relay a signal to the central nervous system about the current state of the body^38–41^. Another fMRI study also employed a similar approach, with subjects attending to heartbeat as well as to skin temperature during the task^38^. Furthermore, numerous imaging studies investigating interoceptive processing across a variety of psychiatric disorders employed tasks presenting disease-related stimuli (for a review see [^42^].)

The Interoceptive Attention Task also involved an exteroceptive attention control condition, during which the word “TARGET” was presented in the center of the screen and randomly switched color from black to a lighter shade of gray, for 500 ms durations. Participants were instructed to focus their attention on the intensity of these color changes and to count the number of times they occurred during the 10-second exteroceptive trial.

One-half of the trials of the interoceptive and exteroceptive conditions were immediately followed immediately by a response period during which subjects rated the intensity of interoceptive sensations (with “1” indicating no sensation, and “7” indicating an extremely strong sensation), or the number of color changes perceived in the exteroceptive trial, via an MRI-compatible button-box. After each rating, there was an intertrial interval (ITI) consisting of a fixation crosshair before the next block begun. The task was performed using E-Prime® 3.0 software (Psychology Software Tools, Pittsburgh, PA, USA).

After receiving verbal instructions about how to perform the task, all subjects underwent a practice session during which they were monitored while making stimulus intensity responses and were asked to indicate whether they had any remaining questions about the task demands.

### Data Analysis

Data processing of fMRI data was performed using AFNI (v16.2.16 [40]; http://afni.nimh.nih.gov/afni). The three runs of the task were processed together. Pre-processing steps are detailed in supplementary material.Maps (βs) for each interoception conditions (body, heart, and stomach) against the exteroception condition (target) were created for each subject. A multivariate modeling using the AFNI program 3dMVM was set for the analysis. We evaluated the main effects of group (FMD or HCs) and interoceptive modality (heart, stomach, or body) and their interaction. For the IA conditions, all subject-level βs represent the signal change from the exteroceptive baseline condition. We performed comparisons across conditions within the same model. Imaging results are reported at a voxel-wise threshold of p<0.005 and cluster size of 20, bi-sided, for a whole brain corrected significance of p <0.05.

To examine the relationship between brain activation during disease-related somatic (body) *vs* visceral interoception (stomach+ heart) and self-report measures of interoception as well as clinical measures of FMD, we correlated the individualβvalues derived from the contrast of somatic versus visceral interoception with the following questionnaire scores: MAIA, HAM-A, HAM-D, and FMDRS, using GraphPad Prism software version 8.0. Correlations were Bonferroni corrected to control for family-wise error for a significance level of *p*□<□0.05.

## Results

### Demographic and clinical characteristics

Twenty-six participants, consisting of 13 patients with FMD and 13 age- and sex-matched HCs, were included in the analysis. Groups did not differ in terms of demographic data and exposure to childhood trauma (Table 1). Compared to HCs, patients reported greater anxiety symptom severity in the seven days prior to the study, which was in the mild range (HAM-A ratings= 10.3 ± 5.1), whereas both patients and HCs did not report depressive symptomatology (score < 10), as assessed by the HAM-D scale. Five patients had a lifetime diagnosis of comorbid psychiatric disorders (depressive disorders n=2, generalized anxiety disorder n=3, PTSD=1).

**Table 1.** Demographic and baseline characteristics of the study sample.

|  | FMD<br>(n=13) | Healthy Controls<br>(n=13) | *p* |
| --- | --- | --- | --- |
| Age – yrs.(sd) | 45.7 (11.6) | 44.5 (10.1) | 0.77 |
| Sex | 10F | 10F | 0.69 |
| HAM-D - mean(sd) | 5.6 (3.3) | 1.2 (1.6) | 0.0002 |
| HAM-A - mean(sd) | 10.3 (5.1) | 1.2 (1.9) | 0.0001 |
| CTQ | 43.6 (14.5) | 38.4 (18.6) | 0.21 |
| Illness Duration - yrs.(sd) | 10.7 (7) | NA | -- |
| FMDRS - mean(sd) | 14.5 (6.8) | NA | -- |
| Comorbid Psychiatric Disorders (n) | 5 | NA | -- |

HAM-D=Hamilton Rating Scale for Depression; HAM-A =Hamilton Anxiety Rating Scale

Clinically, patients reported an average FMD duration of 10.7 years (±7), with a baseline S-FMDRS score of 14.5 (± 6.8). The abnormal movements included tremor (n = 7; seven upper extremities and three lower extremities), dystonia (n = 3 upper limb/shoulder), positive myoclonus (n = 2; shoulder/lower limb); mixed tremor/dystonia (n = 1; upper limb).

### Imaging Results

Whole brain analysis revealed a significant interaction between group (FMD > HCs) and stimulus (interoception□>□exteroception) [*p*< 0.05, corrected] in a large cluster encompassing the left and right posterior cingulate cortex (PCC) and the precuneus, as well as in the left and right caudate nucleus (CN), and in the right anterior insula (aINS) (Figure 1, Table 2). Specifically, compared to controls, patients with FMD exhibited increased hemodynamic response in these regions across interoceptive modalities.

**Figure 1.**
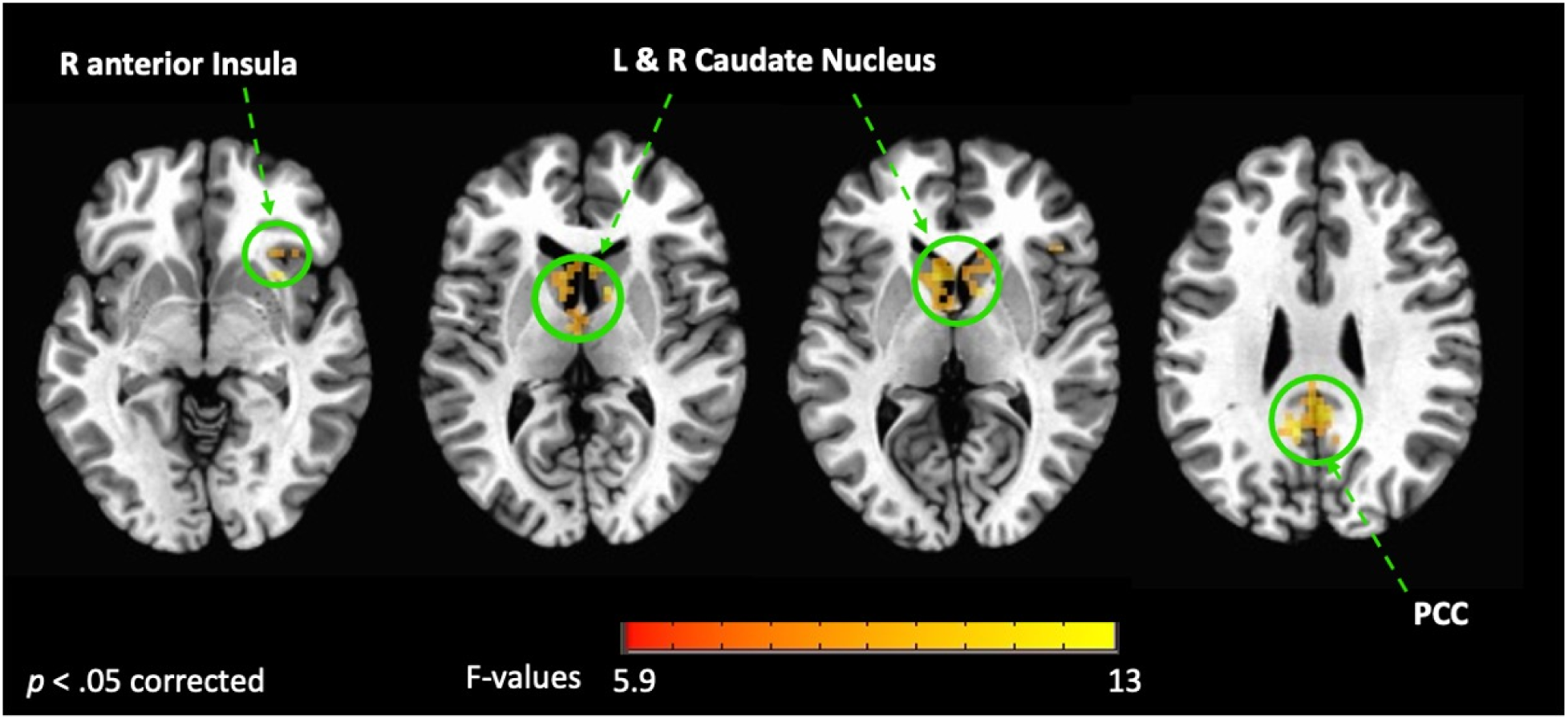
Interaction between groups (FMD vs HCs) and condition (interoception vs exteroception). This figure shows the brain regions activated during interoceptive conditions vs exteroception in patients with FMD compared to healthy controls (HCs). In these regions, hemodynamic activity was increased across interoceptive conditions in the FMD group compared to the HC group. All results shown were corrected for multiple comparisons (*p* < 0.05). FMD= functional movement disorder; PCC= posterior cingulate cortex; R= right; L= left.

**Table 2.** Brain regions exhibiting group-differences in the hemodynamic response to interoception conditions versus exteroceptive attention.

|  |  |  |  |  |  |
| --- | --- | --- | --- | --- | --- |
| *FMD vs Healthy Controls* |  |  |  |  |  |
|  | Side | CM Coordinates |  |  | Voxels |
|  |  | x | y | z |  |
| *Interoception > exteroception (B+S+H > T)* |  |  |  |  |  |
| Posterior Cingulate Cortex | L & R | + 1.5 | + 38.9 | + 30.1 | 112 |
| Caudate Nucleus | L | + 4.8 | + 1.6 | + 8.7 | 77 |
| Insula/ BA13 anterior | R | - 31.9 | - 17.0 | - 2.9 | 26 |
| Caudate Nucleus | R | - 9.4 | - 10.8 | + 5.0 | 25 |
| *Body > Target* |  |  |  |  |  |
| Posterior Cingulate Cortex / Precuneus | L & R | + 0.0 | + 49.3 | + 28.2 | 283 |
| Angular Gyrus | L | + 44.3 | + 61.1 | + 31.5 | 50 |
| Cerebellum (pyramis) | R | - 27.3 | +70.7 | - 31.2 | 31 |
| Middle Frontal Gyrus | L | + 22.3 | -20.8 | +41.5 | 26 |
| Caudate Nucleus | L | +4.8 | -11.3 | +6.0 | 23 |
| Caudate Nucleus | R | - 9.7 | -12.9 | +3.9 | 20 |
| *Stomach > Target* |  |  |  |  |  |
| Precuneus | L & R | -1.5 | +52.5 | +20.5 | 49 |
| *Body > Stomach+Heartbeat* |  |  |  |  |  |
| Posterior Cingulate Cortex | L & R | + 1.5 | + 37.5 | + 26.5 | 175 |
| Thalamus | L | + 1.5 | + 16.5 | + 14.5 | 133 |
| Insula middle | R | - 43.7 | - 4.3 | + 5.6 | 29 |
| Caudate Nucleus | L | + 21.4 | - 15.4 | - 3.0 | 22 |
| Angular Gyrus | L | + 43.3 | + 61 | + 33.9 | 22 |

<sup>a</sup> In all cases, activity was greater in the FMD group
<sup>b</sup> All coordinates reported according to the Talairach stereotaxic atlas computed from template TT\_N27. CM= center mass.

In separate comparisons of each interoception condition (heart [H], stomach [S], body [B]) to the exteroception condition (target [T]), we found significant group differences during somatic interoception compared to the exteroceptive condition (B > T), such that FMD patients had greater average activity in several regions part of the default mode network [DMN] (i.e., posterior cingulate, precuneus, angular gyrus, and medial prefrontal cortex), in the right cerebellum as well as in the bilateral CN [*p*< 0.05, corrected] (Figure 2a; Table 2). Precuneus activity was greater in patients with FMD compared to controls during visceral interoception [*p*< 0.05, corrected] (S > T), whereas no group differences in the hemodynamic response to heartbeat attention were found (Figure 2b; Table 2).

**Figure 2.**
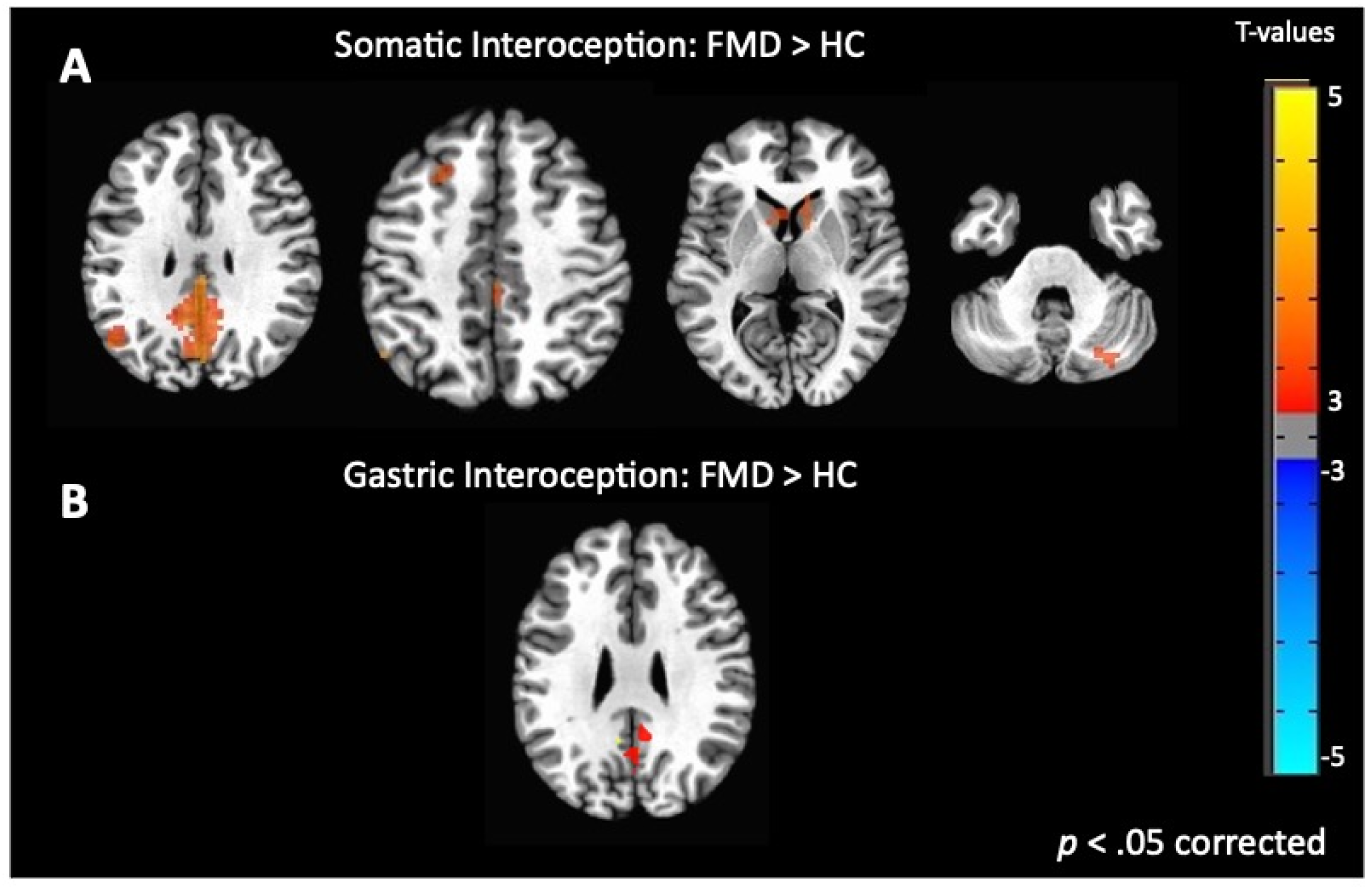
Group differences in somatic and gastric interoception. In the FMD group, attending to disease-related interoceptive signals (A) elicited activity in several regions part of the default mode network, including the bilateral posterior cingulate/precuneus, left angular gyrus, right cerebellum as well as in the bilateral CN. Precuneus activity was also greater during gastric interoception (B) in patients with FMD compared to the HC group. All results shown were corrected for multiple comparisons (p < 0.05). FMD= functional movement disorder.

Next, we examined differences in BOLD activity during somatic interoception compared to the two visceral interoception conditions (B > S + H) between groups. In FMD patients, IA to bodily sensations was associated with increased BOLD response in several nodes of the DMN (i.e., left angular gyrus, left thalamus, and PCC bilaterally), in the right mid-insula and in the CN [*p*< 0.05, corrected] (Figure 3a, Table 2).

**Figure 3.**
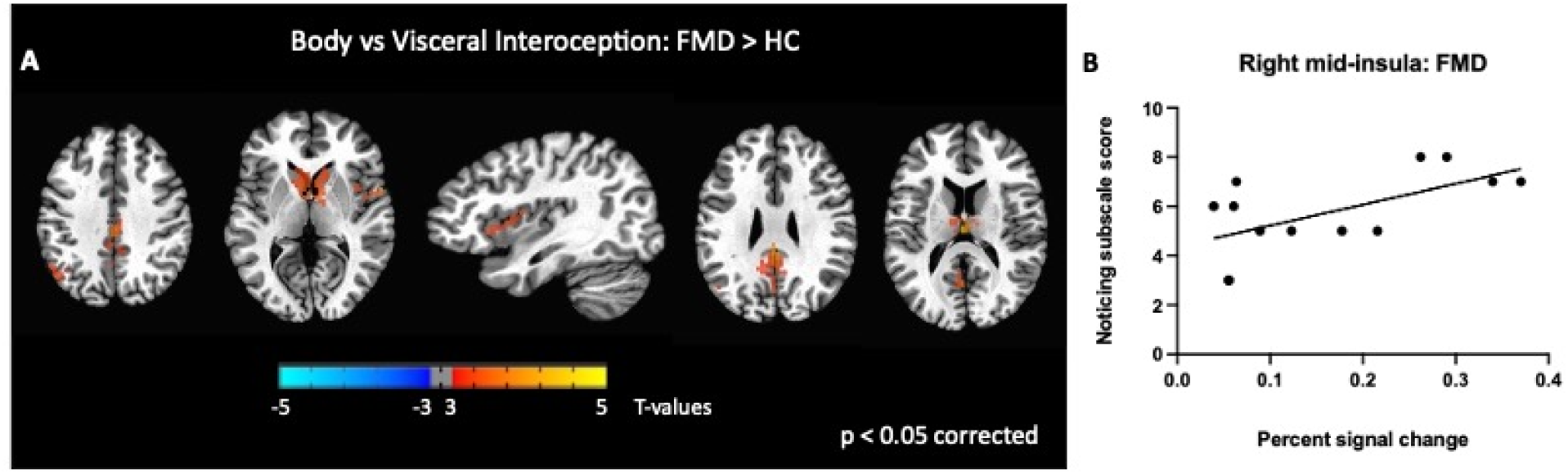
Group differences in somatic, disease-related interoception vs visceral interoception. In the FMD group compared to HCs, attending to disease-related interoceptive signals compared to other interoceptive signals (stomach + heartbeat) (A) elicited activity in several nodes of the DMN (i.e., left angular gyrus, left thalamus, and PCC bilaterally), in the right mid-insula and in the bilateral caudate nucleus. In subjects with FMD, activity in the right mid-insula was positively correlated with the MAIA *Noticing* subscale, which measures body awareness. All results shown were corrected for multiple comparisons ( p < .05). FMD= functional movement disorder.

### Correlations with MAIA scores and with clinical measures of FMD

Scores for each of the eight MAIA subscales did not differ among patients and controls, in line with previous research. In the FMD group, we next assessed the relationship between MAIA subscale scores and BOLD responses in brain regions showing altered response during somatic *vs* visceral interoception (B > S + H). We chose to focus on this comparison, given our hypothesis that disease-related interoceptive signals would hijack interoceptive processing compared to other interoceptive signals.

We found a significant correlation between BOLD activity in the right mid-insula and the MAIA *Noticing* subscale score (r = 0.6; *p*=0.02; Figure 3b). We also examined the relationship between regional BOLD activity and several FMD-related clinical measures and found no significant correlations with S-FMDRS, HAM-D, and HAM-A total scores (*data not shown*).

## Discussion

To our knowledge, this study is the first to investigate the neural correlates of IA in individuals with FMD compared to healthy controls and to examine group-differences between hemodynamic responses to disease-related interoceptive signals *vs* other bodily signals. Our findings provide experimental evidence that FMD is associated with abnormal interoceptive activity in several regions, including the right insula, the bilateral posterior cingulate cortex, and the right caudate nucleus. Group-differences in brain activity were mainly driven by processing of disease-related interoceptive signals, which in patients with FMD was associated with a broader neural activation than monitoring other bodily sensations. Importantly, in this group, activity in the right mid-insula during disease-related *vs* visceral IA was positively correlated with the *Noticing* MAIA subscale scores, which measures individual’s body awareness.

In the comparison of interoception *vs* exteroception, we identified brain regions that have been consistently implicated in interoceptive processing, thus confirming the validity of the imaging paradigm used in our study. In line with our hypothesis, patients with FMD exhibited abnormal insular hemodynamic activity across interoceptive conditions relative to HCs, particularly within the right anterior subregion. The insula is a primary interoceptive cortex involved in salience, prediction, cognition, homeostasis, and self/emotional awareness^40,43^. Accumulating evidence suggests different roles for subregions of the insular cortex during interoception, such that bodily signals are mainly projected to the posterior insula and then transmitted forward along the rostrocaudal axis, integrated with other sensory inputs in the mid-insula, and finally re-represented in the aINS to be consciously available^40,44,45^. As such, the right aINS contributes to the conscious interoceptive experience and has been identified as the main neural substrate of IA^46^. Thus, our finding of greater activity in this subregion, together with evidence of abnormal body-centered attention^7^/23/2024 1:35:00 PM, may suggest that patients with FMD may be hypersensitive to, and perhaps constantly monitoring, interoceptive sensations. In support of this hypothesis, we observed that along with right aINS, interoceptive conditions *vs* exteroception also elicited greater activity within the PCC in FMD participants compared to controls. The PCC is a key node in the DMN^47,48^ and activity in this structure has been associated with decision-making, memory, body ownership, interoceptive and emotion processing, and modulation of arousal state^46,49,50^. Notably, neuroimaging studies have consistently implicated the PCC in controlling the focus (internal *vs* external) and breadth (broad *vs* narrow) of attention, with studies reporting increased PCC activation when individuals direct attention internally^51,52^. Specifically, it has been proposed that the right aINS and the PCC form a system regulating the balance between internally and externally focused attention^49,53^. Thus, the coactivation of PCC and aINS in the FMD group during interoceptive processing *vs* exteroception further suggest that subjects with this disorder may have an impairment in shifting the attentional focus away from interoceptive signals.

Separate comparison of each interoceptive condition *vs* exteroception revealed no group-differences during cardiac interoception, supporting prior reports of normal heartbeat perception accuracy in individuals with FMD^20,21,25,27^. Conversely, increased activity in the precuneus – another key node of the DMN^54^ - was observed during gastric interoception in FMD participants compared to controls, confirming that this network may play an important role in abnormal IA processes in FMD. Furthermore, the precuneus is also implicated in the sense of self and agency^55^, which is impaired in FMD patients^56^, and functional and structural alterations in this region have been reported in association with FMD (for a review see [^57^]). We further observed marked group-differences during processing of disease-related interoceptive signals *vs* exteroception, which in FMD patients elicited a broad activation across the DMN, including the angular gyrus, the mid-frontal gyrus, the PCC and the precuneus. Interestingly, the cerebellum also showed heightened activity in FMD patients compared to controls. This finding is in line with evidence of cerebellar contribution to both attentional^58,59^ and interoceptive processes^60,61^, and can also be explained in the context of prior imaging studies showing increased cerebellar volume and activity in FMD patients compared to either controls or patients with other movement disorders (for a review see [^62^]). Accumulating evidence also suggests that the cerebellum is part of the DMN^63,64^, thus its activation may reflect the widespread engagement of this network observed in our study, particularly in response to disease-related interoceptive signals. Previous studies have found that the DMN is characterized by overactivation and neurometabolic dysfunctions in children and adolescents with FND^65,66^: we expand on these findings by showing that these alterations may represent the neurobiological correlate of abnormal IA in subjects with FMD.

The current study also investigated for the first time group-differences in brain activation during disease-related somatic interoception compared to visceral interoception. As expected, individuals with FMD showed greater BOLD response to disease-related *vs* other bodily sensations in several brain regions. This pattern of activation suggests that ‘disease-centered’ attention allocation may happen not only at the expenses of external stimuli but also of other interoceptive signals. A potential explanation is that these signals are perceived as less salient compared to disease-related sensations, although we did not specifically tested for this hypothesis. However, it is worth mentioning that salience attribution is encoded by the aINS, while our results indicate increased activity in the mid-insula, in line with a prior study in healthy subjects showing that differences based on interoceptive focus (heartbeat *vs* skin temperature) were found in the mid-insula ^11^. According to active inference framework of interoceptive processing, this segment of the insula represents the key neural substrate of interoceptive prediction error, which occurs following a mismatch between interoceptive prediction signals issued by the anterior insula and incoming interoceptive signals arriving via the thalamus^67^. Specifically, the middle and posterior insula compute the difference between the predicted interoceptive signal and the actual interoceptive signal, generating an error signal. Thus, our finding of increased activation in the segment of the insula during disease-related somatic interoception compared to visceral interoception may indicate a greater magnitude of disease-related interoceptive prediction errors, which may induce individuals with FMD to constantly monitor disease-related sensations in the attempt to match them with expected signals. In support of this hypothesis, we did found a positive, although marginally significant correlation between BOLD response in the mid-insula and scores on the *Noticing* MAIA subscale, which assesses the spontaneous tendency to sense or notice bodily sensations.

A predictive coding account of our results is further suggested by evidence of greater activation in several nodes of the DMN, which, together with the insula and other cortical and subcortical regions, are part of unified, intrinsic large-scale brain network, the allostatic-interoceptive network, that modulates visceromotor and interoceptive processes with the goal to maintain or restore allostasis, while also supporting a wide range of psychological functions (emotions, memory, decision-making, pain), which rely on allostasis^60^. Interestingly, this network also includes the CN, which showed greater activation in FMD subjects during disease-related somatic interoception compared to visceral interoception, as well as in the comparison between disease-relate bodily signals and exteroceptive condition. The CN has been associated with perceptual prediction errors^68^, and a study evaluating the neural correlates of breach of expectations during the execution of a sequence of whole-body movements found that prediction-violating movements elicited CN activation^69^. Taken together, our findings suggest that IA, particularly toward disease-related bodily signals, and interoceptive predictive error are closely linked in FMD, with abnormal IA resulting in higher weighting of prediction errors, which in turn may further contribute to maintain the attentional focus on disease-related bodily signals.

No study is without limitations. First, our sample size was modest, although it was in line with previous studies investigating group-differences in behavioral measures of interoception^20,25^ and included cases and controls closely matched for age and sex. Second, concerns about false positive rates in fMRI studies^70^ might be raised given the use of a voxel-wise threshold of p<0.005 and cluster size correction based on random field theory. While recent evidence suggests that these concerns may be overstated^71^, we also believed that the occurrence of false positive rates is unlikely given the extensive amount of neuroimaging studies implicating the insula and nodes of the DMN in interoception. Further, as the current study is the first to investigate the neural correlates of IA in FMD, we decided to adopt a voxel-wise approach and less stringent criteria, to investigate interoceptive relationships across the entire brain. We chose to use a version of the interoceptive task that included monitoring both visceral and somatic signals, in line with a broader conceptualization of interoception as the sensing of signals from the entire body, including skin, muscles, joints, in addition to viscera ^38–41, 72^. This conceptualization is supported by research showing that visceral and somatic afferents converge at several levels on their way to the brain^73^. Furthermore, a study in healthy controls showed that both somatic and visceral interoception activated several brain regions, including the insula, supporting the validity of our study design^11^. However, our characterization of the “heartbeat” and “stomach” conditions as involving visceral interoception and the “body” condition as involving “somatic” interoception involves assumptions that were not directly tested in the current study. As such, interpretation of the differences between interoceptive conditions should take into account the somatic and visceral processing may have been simultaneously engaged during the each interoceptive condition. Finally, we did not employ an accelerometer to record movements during the fMRI scanning, given that previous research suggests that external processes, such as cutaneous sensations, could affect interoceptive processing^41^. However, we acknowledge that these data could have allowed examining the interaction between brain activation and movements evoked by interoceptive attention.

Despite these limitations, this study is the first to unravel the neural correlates of interoception attention in individuals with FMD, highlighting a critical role for regions involved in monitoring body state and in regulating attentional focus and homeostatic inference. Further studies are required to replicate our findings in larger samples and across distinct FND subtypes.

## Supporting information

three runs of the task were processed together. Pre-processing steps are detailed in supplementary material

## Data Availability

All data produced in the present study are available upon reasonable request to the authors

## Acknowledgements

This work has been supported by the NINDS Intramural Research Program and the Mary Ann Tynan Fellowship Program/ Brigham and Women’s Hospital. We also acknowledge the support of the clinical research staff in the NINDS intramural program.

