## Supplementary material for "Functional Movement Disorder Is Associated with Abnormal Interoceptive Brain Activity: A Task-based Functional MRI Study": three runs of the task were processed together. Pre-processing steps are detailed in supplementary material

SUPPLEMENTARY INFORMATION

**fMRI pre-processing**

Preprocessing of fMRI data was performed using AFNI (v16.2.16 [40]; http://afni.nimh.nih.gov/afni). Anatomical images were spatially transformed to the AFNI standard Talaraich space (TT_N27) using the @SSwarper function. Preprocessing of the three runs of task were analyzed together and included removal of spikes, time and special alignment, and non-linear registration. The first two volumes of each voxel’s time course were excluded from analysis to allow the fMRI signal to reach steady state. We set the motion limit at 3 mm and identifying volumes with more than 10% of outliers as defined with 3dToutcount tool in AFNI in a censor file to be used subsequently in the regression analysis. Motion correction and spatial transformation were implemented in a single image transformation. The EPI data were resampled to a 1.75 × 1.75 × 1.75 mm grid and smoothed using a 4 mm full-width at half-maximum Gaussian kernel and using the in-mask option. Each voxel’s mean signal across the time course was then used to normalize the signal value for each EPI volume to percent signal change.
